# Temporal and geographical variation of COVID-19 in-hospital fatality rate in Brazil

**DOI:** 10.1101/2021.02.19.21251949

**Authors:** Tatiana Pineda Portella, Sara Ribeiro Mortara, Rafael Lopes, Andrea Sánchez-Tapia, Maria Rita Donalísio, Marcia C. Castro, Vito Ribeiro Venturieri, Camila Genaro Estevam, Ana Freitas Ribeiro, Renato Mendes Coutinho, Maria Amélia de Sousa Mascena Veras, Paulo Inácio Prado, Roberto André Kraenkel

## Abstract

**Background:** Previous studies have shown that COVID-19 In-Hospital Fatality Rate (IHFR) varies between regions and has been diminishing over time. It is believed that the continuous improvement in the treatment of patients, age group of hospitalized, and the availability of hospital resources might be affecting the temporal and regional variation of IHFR. In this study, we explored how the IHFR varied over time and among age groups and federative states in Brazil. In addition, we also assessed the relationship between hospital structure availability and peaks of IHFR.

**Methods:** A retrospective analysis of all COVID-19 hospitalizations with confirmed outcomes in 22 states between March 01 and September 22, 2020 (n=345,281) was done. We fit GLM binomial models with additive and interaction effects between age groups, epidemiological weeks, and states. We also evaluated the association between the modeled peak of IHFR in each state and the variables of hospital structure using the Spearman rank correlation test.

**Results:** We found that the temporal variation of the IHFR was heterogeneous among the states, and in general it followed the temporal trends in hospitalizations. In addition, the peak of IHFR was higher in states with a smaller number of doctors and intensivists, and in states in which a higher percentage of people relied on the Public Health System (SUS) for medical care.

**Conclusions:** Our results suggest that the pressure over the healthcare system is affecting the temporal trends of IHFR in Brazil.

**Key Messages:**

1. Temporal variation of age adjusted In-Hospital Fatality Rate (IHFR) was markedly heterogeneous among Brazilian states from March to September of 2020.
2. In several states, the IHFR increased in association with the increase in the number of hospitalizations, which suggests that the overload of the healthcare system might be affecting the temporal trends of IHFR in Brazil.
3. The IHFR remained low in the states with higher rates of hospital resources, even with the high demand for hospitalization.
4. The number of doctors and intensivist physicians per habitant was more strongly correlated with the peak of IHFR in the Brazilian states than the number of ICU beds.

## Introduction

COVID-19 has a high and variable in-hospital fatality rate (IHFR), with values ranging from 13% to more than 50%, depending on the region.^1^ It is believed that the disparity in health care accessibility might be affecting the IHFR between regions^2^. This hypothesis has been recently supported by the findings of Ranzani^3^ and Andrade^4^ that national surveillance data in Brazil show higher IHFR in the regions with the lowest number of hospital beds per inhabitant.^3^

New knowledge brought to light by clinical studies and the experience gained by physicians is expected to lead to improvements in the effectiveness of treatment of inpatients with COVID-19 and, consequently, to a reduction of the IHFR over time.^1^ Such a temporal trend has already been reported by Horwitz^5^, who found an 18% reduction of COVID-19 IHFR in a single health system in New York between March and June, and by Dennis^6^, who showed a 20% reduction in England between March and May. Conversely, the IHFR of COVID-19 increased when hospital admissions were higher, even in macroregions where admissions were low at the beginning of the pandemic in Brazil.^3^ This finding highlights the impact of the healthcare system burden in IHFR in a middle income country. However, this trend was not found in all of the five macroregions, nor was adjusted for the possible effects of temporal and regional variation of deaths by age group. Additionally, macroregions in Brazil have little administrative significance compared to the federal states, which have marked differences in the availability of hospital resources^7^ and demographic structure even within the same region. Therefore, the heterogeneity of IHFR trends already found at the region level begs for a finer investigation at the state level.^3^

In order to better understand variations in IHFR over time and between spatial units it is essential to account for age. Several studies have shown that older patients have a higher risk of death by COVID-19 ^2,3,4,8^, and thus a decrease in the IHFR could be attributed to a decrease in hospitalization of elderly people over time. Similarly, the variation in IHFR between regions or states could reflect a difference in the age pattern of hospitalized patients, age structure of the population, in addition to, or rather than a shortage of hospital resources.

By the end of 2020, Brazil was the third country with the highest number of cases of COVID-19 in the world, and the second in number of deaths. The mandatory notification of hospitalized patients with severe acute respiratory infection (SARI), which includes COVID-19, in a nationwide system in Brazil allows us to analyze the variation of IHFR for the entire country during the epidemic. The goal of this study is to assess the effects of time, inpatient age, and states on the IHFR in Brazil. Furthermore, we evaluate the influence of healthcare resources availability on the IHFR in different Brazilian states. Our hypotheses are that (1) IHFR increases with age and decreases over time (as management of severe cases improves and the knowledge is disseminated) but the intensity of this effect varies from state to state; and (2) the IHFR peak is greater in states with worse hospital resources, such as those with a lower number of ICU beds and physicians.

## Methods

### Data

Severe acute respiratory syndrome (SARS) or severe acute respiratory illness (SARI) cases are reported in Brazil in a nation-wide surveillance system, SIVEP-Gripe^9^. SARS/SARI is defined as a flu-like illness with any of the following symptoms: dyspnea (difficulty breathing), persistent chest pressure, O_2_ saturation below 95% in ambient air, bluish color of the lips or face. All hospitalized deaths due to SARS/SARI, regardless of whether the patient was hospitalized or not, enter the SIVEP-GRIPE database.

SIVEP-GRIPE was implemented during the A(H1N1) influenza pandemic in 2009, and since the beginning of COVID-19 pandemic, notification of all cases of SARS/SARI became compulsory. All cases reported in the SIVEP-GRIPE system are tested for etiologic agents and classified as “confirmed SARS-CoV-2”, “influenza”, “cases due to any other etiologic agent”, or “SARS of an unspecified origin”. For the 2020 COVID-19 pandemic, each case notified in SIVEP-GRIPE is tested for SARS-CoV-2, and if confirmed, it is classified based on laboratory results (RT-PCR or serology) and/or clinical or radiological manifestations. These cases are reported on a standardized form, containing patient identification, clinical, laboratory, and epidemiological data.

We extracted data from SIVEP-GRIPE from March 01, 2020 until September 28, 2020 and included all COVID-19 hospitalized cases with a confirmed outcome, according to the above mentioned criteria until epidemiological week 35. We aggregated cases (n=345,281) by epidemiological week of symptom onset, age groups (classified as 0-19, 20-39, 40-59, and 60+ years), state of residence, and outcome (death or discharge). We considered the beginning of the time series as the week in which each state accumulated at least 30 hospitalizations.

### Statistical analysis

#### Generalized Linear Models

To describe the variation in IHFR across epidemiological weeks, states, and age groups, we proposed four alternative generalized linear models (GLM) models: (1) temporal variation in IHFR is related to state characteristics and age pattern of inpatients, (2) temporal variation in IHFR is solely related to the age pattern of inpatients, (3) temporal variation in IHFR is solely related to state characteristics, and (4) temporal variation in IHFR is independent of state and age pattern of inpatients.

With this modelling approach we thus posed competing statistical models for isolated effects of each variable, and their combined effects. We implemented the combination of variables (age group, state, and epidemiological week) as additive and interactive effects in GLM with IHFR as a binomial response variable (number of deaths out of total admissions each week). The time-evolving predictor (an integer value expressing the epidemiological week) was included in the models as a second-order polynomial to allow for non-monotonic evolution of IHRF through time. We adopted an information-based model selection approach to simultaneously confront all linear models that translate each hypothesis outlined above.^10^ We selected the best model based on the Akaike Information Criterion (AIC) and models with ΔAIC < 2 were considered equally plausible.

The analysis included only the 22 Brazilian states that reached more than 1,000 total hospitalizations as of September 28. All data analysis was performed in R^11^ using stats and bblme^12^ packages.

### Spearman Rank Correlation test

To assess the relationship between the IHFR and hospital structure among states we selected the maximum value of IHFR of each state and calculated Spearman correlation with the following variables: (1) number of physicians per 100,000 habitants, (2) number of intensivists care physicians per 100,000 habitants, (3) number of ICU beds (private and public) per 100,000 habitants, and (4) proportion of the population that solely rely on the public healthcare system (SUS) (Table S1). All the analyses were done using R, and data and R scripts are available at https://github.io/covid19br/IHFR.

## Results

Temporal trends in IHFR varied among age groups in different ways across states. Accordingly, the regression model best supported by the data takes into account the additive effects of age group, states, and epidemiological week on IHFR, as well as all second order interactions between these effects (Table 1, Figure 1). Amazonas (AM) state exhibited the highest estimate of IHFR (Figure 1 A) and Minas Gerais (MG), the lowest IHFR (Figure 1 V). IHRF was over 70% in five states (Figure 1 A-E), between 70 and 50% in nine states (Figure 1 F-N), and under 50% only in eight states (Figure 1 0-V). In all states that showed a decrease in IHFR over time, the peak of IHFR was very high or high (Figure 1 A-L).

**Table 1.** Results of the model selection ordered by ΔAICc indicating the best model that fits the data.

| **Model** | **AICc** | **$\Delta AICc$** | **df** | **Akaike weight** |
| --- | --- | --- | --- | --- |
| Week * Age +<br>Week * UF | 9919.83 | 0 | 138 | 1 |
| Week + Age +<br>UF | 11188.29 | 1268.46 | 27 | <0.0001 |
| Week * Age | 17698.14 | 7778.31 | 12 | <0.0001 |
| Week + Age | 17735.80 | 7815.97 | 6 | <0.0001 |
| Week * UF | 52305.68 | 42385.85 | 66 | <0.0001 |
| Week + UF | 53702.73 | 43782.90 | 24 | <0.0001 |
| Week | 61856.93 | 51937.10 | 3 | <0.0001 |

**Figure 1.**
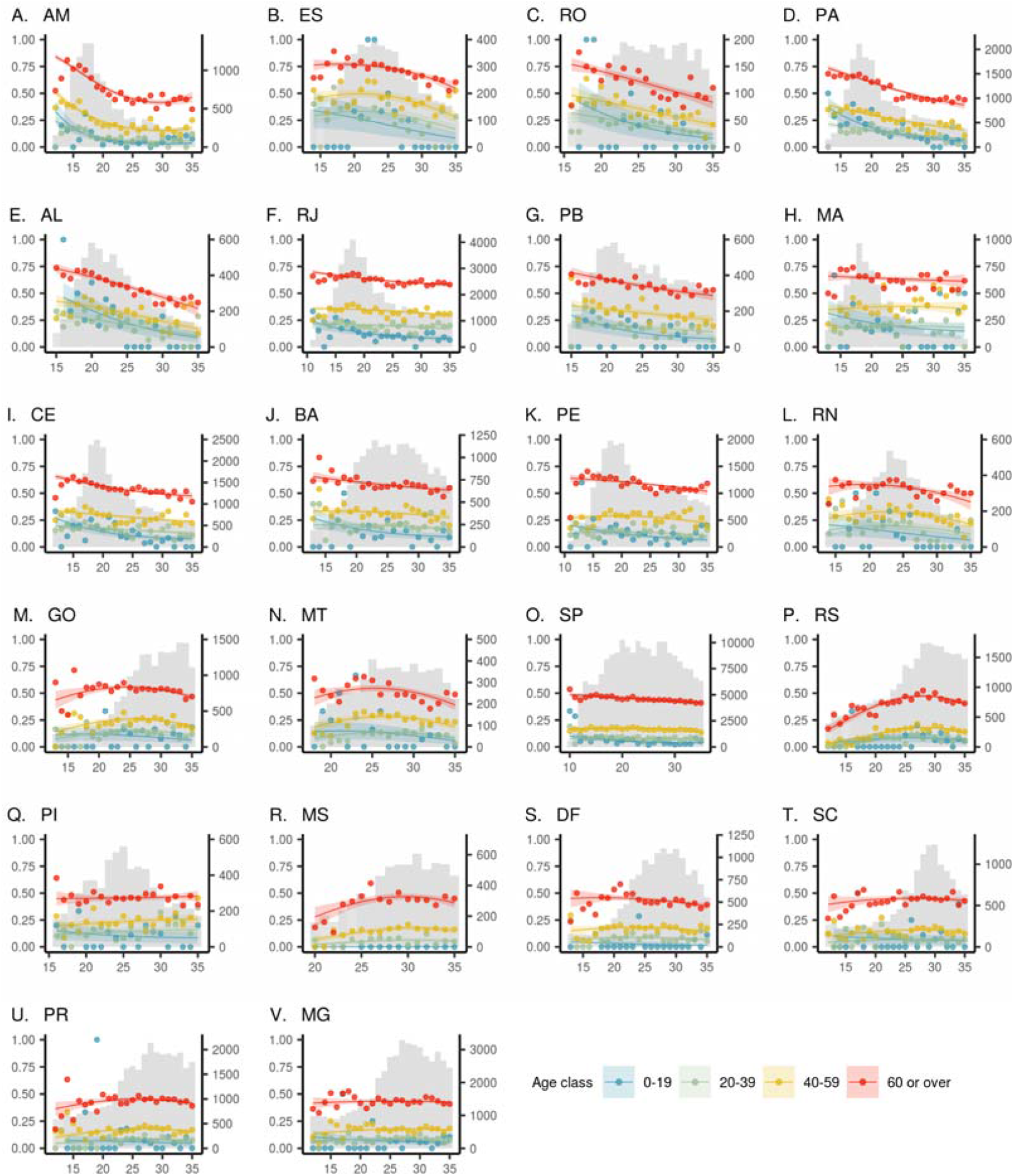
Temporal variation on In-Hospital-Fatality Rates of COVID-19 by age group and states of Brazil. States are ordered from the highest estimated peak of IHFR to the lowest. Dots represent the observed IHFR values, curves represent the IHFR estimated by the model per age group, and shaded area, the confidence interval of estimates. Grey bars on the second y-axis indicate the temporal variation on SARS hospitalizations. The geographic location of 22 states analyzed in Brazil is in Figure 2A. AM = Amazonas, ES = Espírito Santo, RO = Rondônia, PA = Pará, AL = Alagoas, RJ = Rio de Janeiro, PB = Paraíba, MA = Maranhão, CE = Ceará, BA = Bahia, PE= Pernambuco, RN = Rio Grande do Norte, GO = Goiás, MT = Mato Grosso, SP = São Paulo, RS = Rio Grande do Sul, PI = Piauí, MS = Mato Grosso do Sul, DF = Distrito Federal, SC = Santa Catarina, PR = Paraná, MG = Minas Gerais.

In all the states and epidemiological weeks, IHFR was higher in patients aged 60 and over (ranging from 16-84%), followed by patients aged 40-59 (3-50%). The IHFR of patients aged 20-39 (1-33%) was higher than those in the age group of 0-19 years (0-46%) during most of the epidemiological weeks in seven states from three regions: RJ, CE, BA, PE, SP, MS, DF (Figure 1 F, I, J, K, O, R, S). In the remaining states, the IHFR of patients aged 0-19 and 20-39 was the same during most of the studied period (Figure 1).

The IHFR of patients aged 60 years and over decreased over time in 14 states, and in most of them this reduction accompanied the decreasing trend of their hospitalizations. This temporal decrease in IHFR was more accentuated in states in which the IHFR was initially higher (> 70%) and with more limited health care structure (Figure 2, all states in North, most states in Northeastern and two states in Southeastern region). In only four states (BA, SP, DF, and MG) the estimated IHFR decreased while hospitalizations increased or were stable (Figure 1. J, O, S, V). Additionally, in eight states the IHFR of the elderly group increased over time. This increasing trend was in line with a later increase of hospitalizations, notably in the central and southern regions. Among most of the states that experienced a late increase of IHFR or even a decrease despite increasing hospital admittances, the initial IHFR was the lowest (<50%) and did not exceed 50% at any time. Within each state, the temporal trend of IHFR for age groups 20-39 and 40-59 was similar to the trend of patients aged 60 years and over, and the temporal trend of patients aged 0-19 diverged from the other age groups, decreasing slightly over time in most of states (20 of 22).

**Figure 2.**
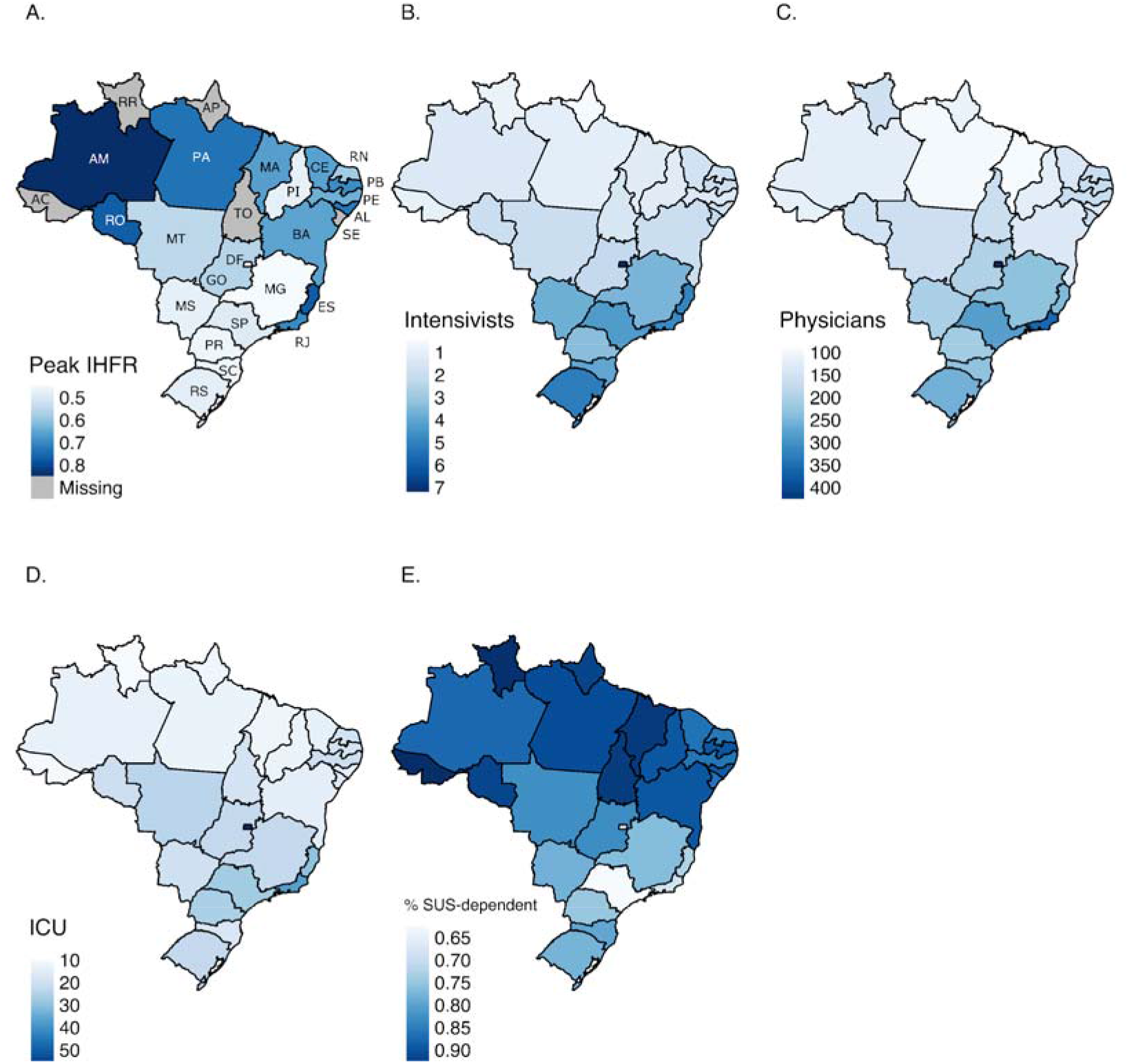
(A) Predicted peak of IHFR, (B) Number of intensivists, (C) physicians, (D) ICU beds per 100.000 habitants, and (E) percentage of the population depending on the SUS at the state level in Brazil.

The lowest IHFR peaks for patients aged 60 years and over (Figure 2 A) were found in the states with the highest numbers of intensivists (Figure 2 B), physicians (Figure 2 C), and ICU beds per habitant (Figure 2 D) and in the states with the lowest proportion of SUS-dependent citizens (Figure 2 E). In addition, the Spearman correlation test showed a significant (p<0.05) negative correlation between the peak value of IHFR and number of physicians and intensivists (Figure 3 A-B), and a positive correlation (p<0.05) between the peak value of IHFR and proportion of SUS-dependent citizens (Figure 3 D). ICU beds showed no relationship with the peak of IHFR (Figure 3 C).

**Figure 3.**
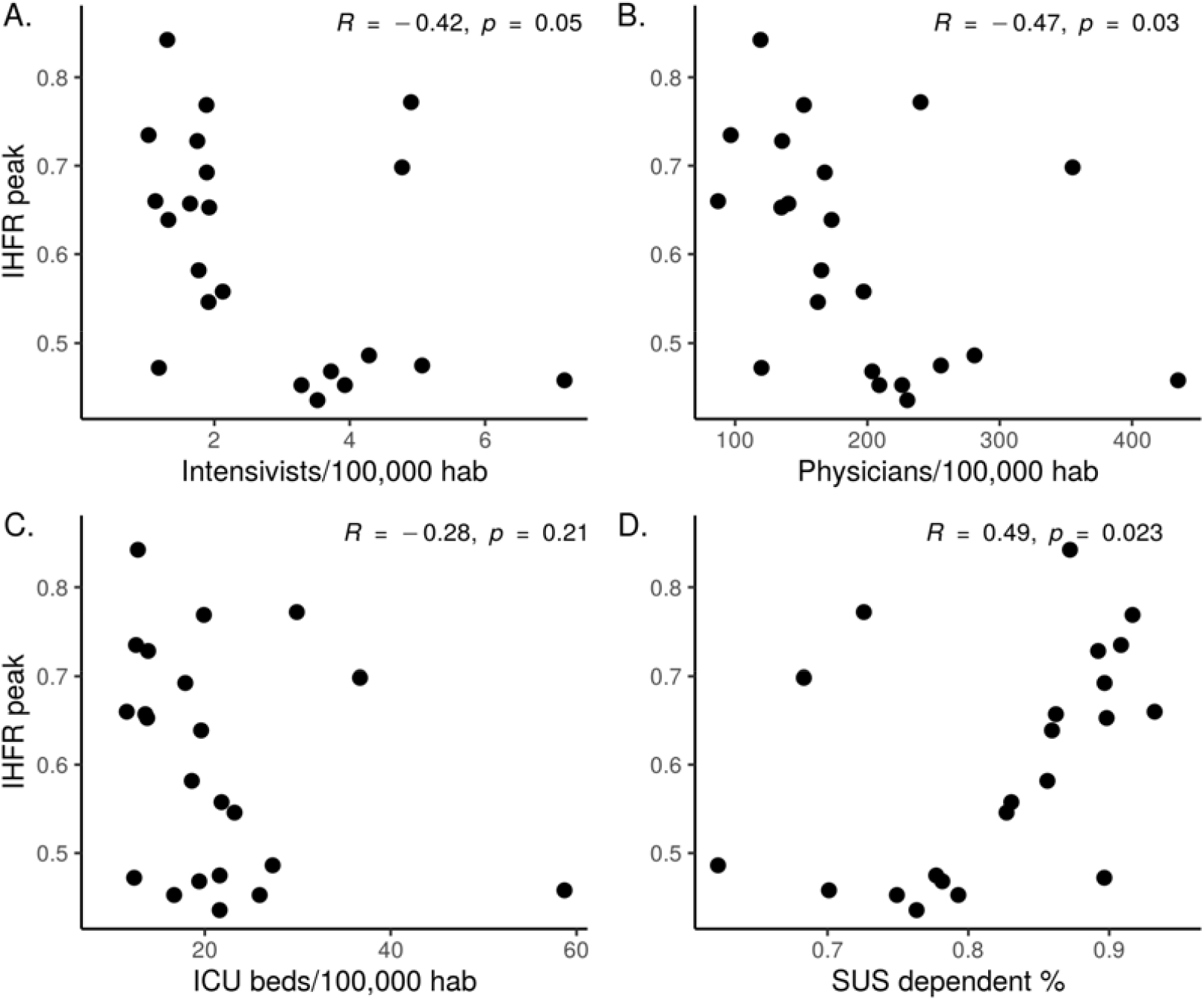
Scatter plot and Spearman Rank Correlation test between IHFR peak of each state and the health indicators: (A) number of intensivists per 100,000 habitants, (B) number physicians per 100,000 habitants, (C) ICU beds per 100,000 habitants, and (D) % of population SUS dependent.

## Discussion

In this study, we showed temporal trends in IHFR, which were markedly heterogeneous among Brazilian states during the study period. In addition, IHFR peaks along these trends were higher in states with fewer health care resources, especially those with less doctors and intensivists per 100,000 inhabitants, which were also the states in which a higher percentage of people relied on public health services (SUS) for medical assistance.

The patterns of temporal variation of the IHFR for some of the Brazilian states reported in this study coincide with the ones for macroregions reported in Ranzani^3^ and were partly different from a declining trend that has been reported for the majority of countries so far.^1,5,6^ It has been hypothesized that the decrease in temporal variation in the IHFR might be related to a better management of patients, due to the knowledge and experience gained by physicians over time, for instance incorporating anticoagulants^13^, corticosteroids in severe cases^14^, more accurate indication of mechanical ventilation, and respiratory physiotherapy.^15,16^. However, our results show that in some Brazilian states the IHFR per age group increased during the study period. Thus we hypothesize that, in addition to the experience gained by the health personnel, other factors might be affecting the temporal variation in the IHFR.

The IHFR peaked during the weeks with the highest number of hospitalizations in most states, suggesting that hospital overwhelming is affecting COVID-19 inpatients mortality. This hypothesis was also raised by Ranzani^2^, however. They didn’t consider possible effects of the variation in the deaths by age group in the IHFR. Although data on hospital bed occupancy over time is unavailable for the Brazilian states, Andrade^17^ reported that in several regions of Brazil hospitals were operating near full capacity during the most critical moments of the pandemic. The impact on the increase in-hospital mortality has been described in epidemic periods (influenza) and increased occupancy of ICU beds.^18,19^. However, our results suggest that some states with higher rates of hospital resources were able to avoid the collapse of the system during increased demand for hospitalization. This effect could be observed especially in SP, BA, MG, and DF where the hospital lethality remained low and still decreased with higher rates and/or an increase in the number of hospitalizations.

Previous studies suggested that the high bed occupancy rates and/or high patient volumes in the emergency department were correlated with the increase in the IHFR due to a deterioration in the quality of medical care ^18,19,20^ or to a bias in the selection of patients with worse clinical conditions, which therefore are more likely to die.^20^ However, the increase in lethality rates in the Brazilian states could also have been caused by the overload of their healthcare network. In Brazil, ill patients generally pass through a triage before being sent to a healthcare facility. In general, the most critically ill patients are sent to high complexity hospitals, which are usually located in large urban areas, having high technological resources and specialized teams that can provide more qualified clinical support to these patients.^21^ However, with the increase of COVID-19 cases, many of these hospitals could have achieved their maximum capacity, causing many COVID-19 severely ill patients to be sent to smaller and less complex hospitals with little or no specialized health care staff. This could have contributed to the increase in the number of COVID-19 IHFR during the higher burden of hospitalizations, especially in the states that had already been suffering from a lack of hospital resources.

Ranzani^3^ also found the highest crude IHFR in the macroregions of Brazil with the lowest number of hospital and ICU beds. However, our study suggests that the lack of doctors and intensivists is more important to the higher IHFR than the number of ICU beds. Although huge efforts have been made to increase the number of hospital beds and artificial ventilators in Brazil since the beginning of the pandemic, no similar efforts were made to increase the number of physicians. Brazil has a ratio of physicians per 100,000 inhabitants comparable to some developed countries^22^, but most of them are based in south and southeast regions and few professionals have been willing to work in other regions, especially outside the capital cities.^23,24^ In addition, Brazil still suffers from a shortage of medical specialists, where in 2017 only 62,5% of physicians have medical residency training.^25^ Thus, it is likely that many hospitals that were able to expand beds during the pandemic may have not been successful in expanding the medical staff, especially those specialized in intensive care, which is among the most relevant specialties for the management of severe cases of COVID-19. In addition, the states with fewer doctors and intensivists have also a greater dependence on SUS for care, which is usually the only alternative for health assistance for the lower socioeconomic strata of the population. SUS has been severely underfunded due to the Brazilian economic crisis^26^, which also has affected the mortality rates of the most vulnerable populations.^27,28^ Particularly in a time of health crisis such as the COVID-19 pandemic, SUS dependency exposed and intensified the social inequity of the most vulnerable population.

We acknowledge some limitations in our study. First, we use routinely collected secondary data on hospitalized cases of severe acute respiratory syndrome and deaths. While these data may carry some uncertainty, particularly in periods of high hospital demand and peak incidence, they are the best available official data and they are used to guide policy and decision making. Second, state analysis does not capture idiosyncrasies observed among the municipalities within the states, so even more variability is expected at the sub-state level. The testing capacity and quality of epidemiological surveillance are heterogeneous across states, with different levels of expertise for investigating and reporting cases. Lastly, our models do not include other socioeconomic factors that could affect the IHFR, such as variations in prevalence of comorbidities and nutrition status.

To the best of our knowledge, this study presents the first panel of temporal trends of in-hospital fatality rate accounting for age groups in Brazilian states, and contributes to the understanding of regional disparities revealing the mosaic of epidemics of COVID-19 in the country. Contrary to previous studies that found a reduction of the IHFR of COVID-19 over time ^2,5,6^, our results show great heterogeneity in the IHFR temporal trends. In several states, the IHFR increased in association with the increase in the number of hospitalizations, which suggests that, in addition to medical learning, the overload of the healthcare system might be affecting the temporal trend of IHFR in Brazil. In this study, we also found that the highest lethality peaks occurred in states with lower availability of hospital resources, especially doctors and intensivists physicians. In particular, this result highlights the importance of the health care structure, not only to ensure care for all severely ill patients but also to increase the chances of survival of those who are already hospitalized.

## Supporting information

Source of the state-level hospital structure variables.

## Data Availability

All data used are publicly available. The raw data, dictionary, and code used for the analysis are available at github.io/covid19br/IHFR

https://github.com/covid19br/IHFR_manuscript

## Funding

This work was partially supported by the Coordenação de Aperfeiçoamento de Pessoal de Nível Superior - Brazil (Finance Code 001 to TPP, AST, CGE), Conselho Nacional de Desenvolvimento Científico e Tecnológico – Brazil (grant number: 141698/2018-7 to RL, 313055/2020-3 to PIP, 312559/2020-8 to MASMV, 311832/2017-2 to RAK) and Fundação de Amparo à Pesquisa do Estado de São Paulo-Brazil (contract number: 2016/01343-7 to RAK).

## Acknowledgments

We are very thankful to all researchers of Observatório COVID19 BR Team for their support and hard work during the studies of the COVID-19 progress in Brazil.

## Data availability

All data used are publicly available. The raw data, dictionary, and code used for the analysis are available at https://github.com/covid19br/IHFR_manuscript

## Conflict of interest

We declare no competing interests

## Author Contributions

TPP, RAK, PIP, RMC, SRM, RL and AST conceived the study and its design with subsequent inputs from all authors. TPP, SRM, AST, RL and CGE organized and entered data. TPP, SRM, PIP, RMC, RL and AST contributed to data analyses. TPP, SRM, AST and RL had access to the underlying data for verification. TPP, PIP, MRD, MASMV, VRV, AFR, MCC, CGE, RMC, SRM, AST, RL and RAK contributed to data interpretation. TPP, RL, MRD, VRV, SRM, PIP and MCC wrote the main draft of the manuscript with input from all the authors. All authors contributed to the final drafting of the manuscript.

## Notes

### Competing Interest Statement

The authors have declared no competing interest.

### Funding Statement

This work was partially supported by the Coordenacao de Aperfeicoamento de Pessoal de Nivel Superior, Brazil (Finance Code 001 to TPP, AST, CGE), Conselho Nacional de Desenvolvimento Cientifico e Tecnologico, Brazil (grant number: 141698/2018-7 to RL, 313055/2020-3 to PIP, 312559/2020-8 to MASMV, 311832/2017-2 to RAK) and Fundacao de Amparo a Pesquisa do Estado de Sao Paulo, Brazil (contract number: 2016/01343-7 to RAK).

