## Supplementary material for "Temporal and geographical variation of COVID-19 in-hospital fatality rate in Brazil": Source of the state-level hospital structure variables.

**Support Information**

S1 - Source of the state-level hospital structure variables.

| **Variable** | **Source** | **Link** |
| --- | --- | --- |
| Number of Physicians per 100.000 per State | Medical Demography in Brazil, 2018 | https://jornal.usp.br/wp-content/uploads/DemografiaMedica2018.pdf |
| Number of Intensivists per 100.000 per State | Medical Demography in Brazil, 2018 | https://jornal.usp.br/wp-content/uploads/DemografiaMedica2018.pdf |
| Number of ICU beds per 100.000 per State | Federal Council of Medicine, Brazil | <https://medicinasa.com.br/numero-leitos-uti/> |
| Proportion of public healthcare dependents | National Agency of Supplemental Health (ANS) | <http://www.ans.gov.br/> |
